# Interpreting vaccine efficacy trial results for infection and transmission

**DOI:** 10.1101/2021.02.25.21252415

**Authors:** Marc Lipsitch, Rebecca Kahn

**Author notes:** correspondence to, 677 Huntington Ave, Suite 506, Boston, MA 02115.

## Abstract

Randomized controlled trials (RCTs) have shown high efficacy of multiple vaccines against SARS-CoV-2 disease (COVID-19), and recent studies have shown the vaccines are also effective against infection. Evidence for the effect of each of these vaccines on ability to transmit the virus is also beginning to emerge. We describe an approach to estimate these vaccines’ effects on viral positivity, a prevalence measure which under the reasonable assumption that vaccinated individuals who become infected are no more infectious than unvaccinated individuals forms a lower bound on efficacy against transmission. Specifically, we recommend separate analysis of positive tests triggered by symptoms (usually the primary outcome) and cross-sectional prevalence of positive tests obtained regardless of symptoms. The odds ratio of carriage for vaccine vs. placebo provides an unbiased estimate of vaccine effectiveness against viral positivity, under certain assumptions, and we show through simulations that likely departures from these assumptions will only modestly bias this estimate. Applying this approach to published data from the RCT of the Moderna vaccine, we estimate that one dose of vaccine reduces the potential for transmission by at least 61%, possibly considerably more. We describe how these approaches can be translated into observational studies of vaccine effectiveness.

**Highlights:**

- SARS-CoV-2 vaccine trials did not directly estimate vaccine efficacy against transmission.
- We describe an approach to estimate a lower bound of vaccine efficacy against transmission.
- We estimate one dose of the Moderna vaccine reduces the potential for transmission by at least 61%.
- We recommend separate analysis of tests triggered by symptoms vs. cross-sectional tests.

## INTRODUCTION

Randomized controlled trials (RCTs) have shown high efficacy of multiple vaccines against SARS-CoV-2 disease (COVID-19) [1–6], and recent studies have shown the vaccines are also effective against infection [7,8]. Evidence for the effect of each of these vaccines on ability to transmit the virus is also beginning to emerge [9–11].

It is important to understand the effect of vaccination on infection, shedding and transmission of the virus [12]. This information can inform personal decisions about resuming contact once one has been vaccinated (or one’s contact has), prioritization decisions [13], models of the effect of vaccination [14], and policy [15].

Hypothetically, it is possible that the 60-95% protection offered by these vaccines [1–5,16] against symptomatic disease could (i) be purely protection against symptoms with no effect on infection or transmission (or increased transmission due to decreased case detection), (ii) be largely or entirely due to protection against infection, suggesting an effect on transmission similar to the efficacy against symptomatic infection; or (iii) be 70-95% protective against infection and moreover reduce the shedding of virus by those who do become infected, in which case protection against transmission could be even greater than that against symptomatic disease. The primary endpoint of RCTs to date, however, sheds little light on the magnitude of protection the vaccines could offer against transmission.

The effect of a vaccine on transmission is a composite of its effect on becoming infected (because someone not infected cannot transmit) and its effect on the infectiousness of those who get infected despite vaccination [11]: these components have been called the vaccine efficacy for susceptibility to infection and vaccine efficacy for infectiousness [17]. Under plausible assumptions, the efficacy of a vaccine in preventing transmission can be defined as:

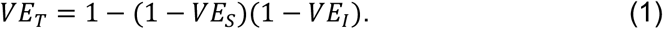

where *VE*_*S*_ and *VE*_*I*_ are the vaccine efficacy against susceptibility (acquiring viral infection) and against infectiousness, respectively [17,18].

RCTs of the Moderna, Astra-Zeneca, and Janssen vaccines have provided some evidence about vaccine effects on the probability that a trial participant will harbor detectable virus by swabbing participants irrespective of symptoms at one or more time points during the trial and testing the swabs by RT-PCR to detect virus [2,16,19]. News reports indicate that those still in placebo-controlled trials will provide ongoing samples that can yield similar data over time [20]. In each case, reduced prevalence of viral positivity in vaccine vs. placebo recipients may be interpreted as a reduction in acquisition or duration or both, with potentially direct relevance to transmission. However, some reports from the original trials present composite measures involving infections detected by screening of non-symptomatic individuals combined with those detected by swabbing of symptomatic individuals [6].

Here we describe the results of simulations of randomized trials that are designed to clarify what information is gained by swabbing individuals for viral infection, how this relates to other measures of vaccine efficacy, and what information is present in measures combining different reasons for sampling (no symptoms vs. symptoms).

We first show that the vaccine effect on viral positivity (*VE*_*V*_) in individuals swabbed at random, regardless of symptoms (i.e. the prevalence odds of those testing positive for viral RNA (equation 4)), is closely approximated by a vaccine efficacy measure previously defined for bacterial carriage, despite several departures from the assumptions underlying the prior work [21]. This measure captures the product of the vaccine’s efficacy in reducing acquisition and its effect in shortening infection duration. We describe how these departures affect the estimates under varying trial conditions. We show that under the reasonable assumption that vaccinated individuals who become infected are no more infectious during their duration of viral shedding than unvaccinated individuals, *VE*_*V*_ is a lower bound on the vaccine’s efficacy against transmission. We recommend that samples taken to assess vaccine effects on viral positivity be taken in a cross-section of the population (or, alternatively, in a cohort of contacts), irrespective of symptoms, and that this outcome be analyzed separately from the outcome of a positive test where the test was triggered by symptoms (the primary endpoint in most RCTs for SARS-CoV-2 vaccines).

## METHODS

We simulate follow-up of 100,000 individuals for 300 days. For each person each day, we conduct a Bernoulli trial to determine if they will be infected that day, with a probability based on an external force of infection. We assume in our baseline simulations that this daily probability remains constant at 0.001 and also examine a higher force of infection of 0.003 in a sensitivity analysis. To examine the effects of deviations from the steady state assumption described in prior work for estimating *VE*_*V*_ [21], we compare scenarios in which individuals immediately become susceptible again after recovery (“SEIS”) to scenarios in which prior infection confers full protective immunity for the duration of follow-up (“SEIR”).

We vary the proportion of cases that are symptomatic (Table S1), with symptom onset occurring five days after infection [22]. After a three day latent period [23], infected individuals shed virus for a period drawn from a uniform distribution of 15-21 days [24,25]. While test sensitivity varies by day of infection [26], we make the simplifying assumption that individuals will test positive on any day they are shedding virus.

On day 100, after the population has reached a pseudo steady state for prevalence, we randomize half of the individuals to receive a two-dose vaccine, with the doses given 28 days apart. We conservatively assume the vaccine confers 50% of its full two-dose efficacy after the first dose and that there is a seven day delay after each dose for immunity to take effect. We model three types of vaccine efficacy (Table S1). First, the vaccine multiplies the probability of infection each day by a factor 1 – *VE*_*S*._ Second, the vaccine multiplies the duration of shedding by a factor 1 – *VE*_*D*_. Third, the vaccine multiplies progression to symptoms among those infected by a factor 1 – *VE*_*P*_. We calculate *VE*_*P*_ based on the value of *VE*_*S*_ and the assumption that the vaccine reduces symptomatic disease by 95% (*VE*_*SP*_) [1,2], using the equation:

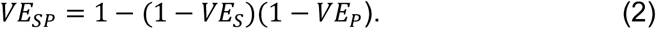

We then simulate testing and estimation of three measures of vaccine efficacy:

### Vaccine efficacy for viral positivity (VE_V_)

We assume all individuals are tested regardless of symptoms on day *t*. Those who are shedding virus on day *t* are counted as positive (i.e. perfect test sensitivity and specificity). We then calculate 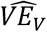 using the prevalence odds ratio comparing vaccinated to unvaccinated using eq. 6 below.

### Vaccine efficacy for non-symptomatic infection (VE_non-symptomatic_)

We estimate vaccine efficacy for non-symptomatic infection by calculating the prevalence odds ratio of PCR positivity among individuals who are not symptomatic on day in the vaccinated vs. unvaccinated groups.

### Vaccine efficacy estimated from a combination of symptoms and routine tests (VE_Combined_)

For this measure of vaccine efficacy, we count as positive any individuals who test positive on day *t* in cross-sectional testing as well as those who were symptomatic and tested positive on or before day t. We then calculate the odds ratio comparing vaccinated to unvaccinated.

Code is available: https://github.com/rek160/InterpretingVaccineEfficacy.

## RESULTS

Table 1 summarizes the notation for various measures of vaccine efficacy studied here.

**Table 1.**

| VE measure | Efficacy against: |
| --- | --- |
| $VE_S$ | Susceptibility to infection (vaccinated person's reduced risk of becoming infected) |
| $VE_I$ | Infectiousness (vaccinated person's reduced probability of infecting others, if they do become infected) |
| $VE_D$ | Duration of shedding (vaccinated person's reduced time of shedding positive if they do become infected) |
| $VE_P$ | Progression to symptoms (vaccinated person's reduced risk of becoming symptomatic if they do become infected) |
| $VE_{SP}$ | Symptomatic disease (vaccinated person's reduced risk of acquiring symptomatic infection, incorporating $VE_S$ and $VE_P$) |
| $VE_V$ | Viral prevalence (vaccinated person's reduced risk of harboring virus at a point in time, incorporating $VE_S$ and $VE_D$) |
| $VE_T$ | Transmission (vaccinated person's reduced risk of transmitting infection, incorporating $VE_S$ and $VE_I$) |
| $VE_{combined}$ | Combined symptomatic incidence & asymptomatic prevalence (vaccinated person's reduced risk of infection in a combined sampling method) |

### For a vaccine that reduces both incidence and duration of viral carriage, vaccine efficacy against carriage can be interpreted as the product of these two effects

Prior work (concerning a bacterial pathogen, though in this exposition we refer to the pathogen as virus) showed that under certain assumptions, for a vaccine that reduces incidence but not duration of infection, the reduction in incidence rate caused by the vaccine (termed in the original paper the vaccine efficacy against acquisition [21], but which we call vaccine efficacy against susceptibility to infection, for consistency with most of the literature [17]) can be defined as

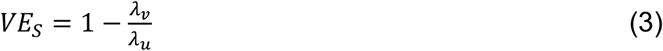

where *λ*_*v*_ is the incidence rate in the vaccinated and *λ*_*u*_ is the incidence rate in the placebo arm, and can be estimated as

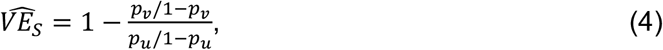

where *p*_*u*_ and *p*_*v*_ are the prevalence of the virus in the placebo and vaccine arm respectively, so the estimator is just one minus the odds ratio for carrying the pathogen for vaccine vs. placebo recipients. As shown by [21], if the vaccine does reduce duration of detectable infection, the quantity

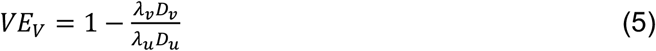

-- vaccine efficacy for viral positivity -- can be defined as the combined effect of the vaccine on incidence and duration, and can be estimated identically

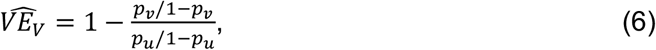

again from the prevalence odds ratio for the vaccine, and now with effects on both duration and incidence, we have the algebraic relationship

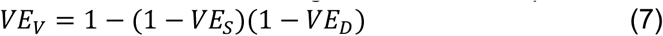

where *VE*_*V*_ is defined in eq. 5, *VE*_*S*_ is defined in eq. 3, and 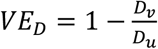 is the reduction in average duration of viral positivity due to the vaccine. Thus, to generalize from reference [21], *VE*_*V*_ is an upper bound on *VE*_*S*_ : *VE*_*V*_ ≥ *VE*_*S*_ with equality for the special case where *VE*_*D*_ = 0.

### *VE*_*V*_ is a lower bound on the vaccine’s efficacy against transmission, assuming that vaccinated individuals who become infected are no more infectious than unvaccinated individuals

Equations 1 and 7 show that *VE*_*V*_ and *VE*_*T*_ are similar though not identical; in particular, they differ in only one term: the substitution of *VE*_*D*_ in the definition of *VE*_*V*_ as opposed to *VE*_*1*_ in the definition of *VE*_*T*_. If vaccinated and unvaccinated virus-positive individuals contributed equally to the force of infection, then these two terms would be identical, and we would have *VE*_*V*_ = *VE*_*T*_ : that is, the reduction in transmission thanks to the vaccine would be the combination of reduced probability of infection and reduced duration of shedding in those infected despite vaccination. If we assume that for every day of being virus positive, a vaccinated infected person is on average no more infectious (and perhaps less due to lower viral loads) than an unvaccinated infected person with the same exposure, then we can conclude that

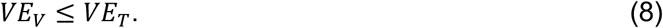

Under this plausible assumption,*VE*_*T*_, which cannot be directly estimated from available trial data, is at least as large as *VE*_*V*_, which can. We therefore proceed to discuss how to estimate *VE*_*V*_ for SARS-CoV-2 vaccines.

### Simulated trials show that these estimators applied to a single cross-sectional swab approximately recover the simulated effects of a vaccine on viral positivity, incorporating effects on acquisition and duration, with visible downward bias just after and long after vaccines are administered

Fig. 1 shows results of 300-day simulations of a trial of 100,000 participants randomized 1:1 to vaccine or placebo on day 100. These participants have been exposed to a constant incidence of infection since day 0. The different panels represent (left to right) simulations with *VE*_*S*_ = 0,0.3,0.6,0.9 and (top to bottom) *VE*_*D*_ = 0,0.3,0.6,0.9. We simulate a 2-dose regimen, 28 days apart with the first dose giving half the full efficacy and the effect of each dose starting one week after it is given, that is, on days 107 and 135 of the simulation. Given the five day incubation period, the vaccine’s effects on symptomatic infection will be observed beginning 12 days after the vaccine dose. The solid black lines give the dose-1 and dose-2 predicted values for based *VE*_*V*_ on eq. 7, while the curves show the estimates 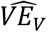 obtained from the simulated data using eq. 6. Fig. 1A shows the situation under the assumption that individuals naturally infected who recover (clear infection) become once again susceptible to reinfection. This is unrealistic for SARS-CoV-2 but follows the assumptions made in the above equations following [21]. Fig. 1B makes the opposite assumption, that individuals naturally infected (whatever their vaccine status) are completely protected against reinfection for the duration of the simulation.

**Figure 1.**
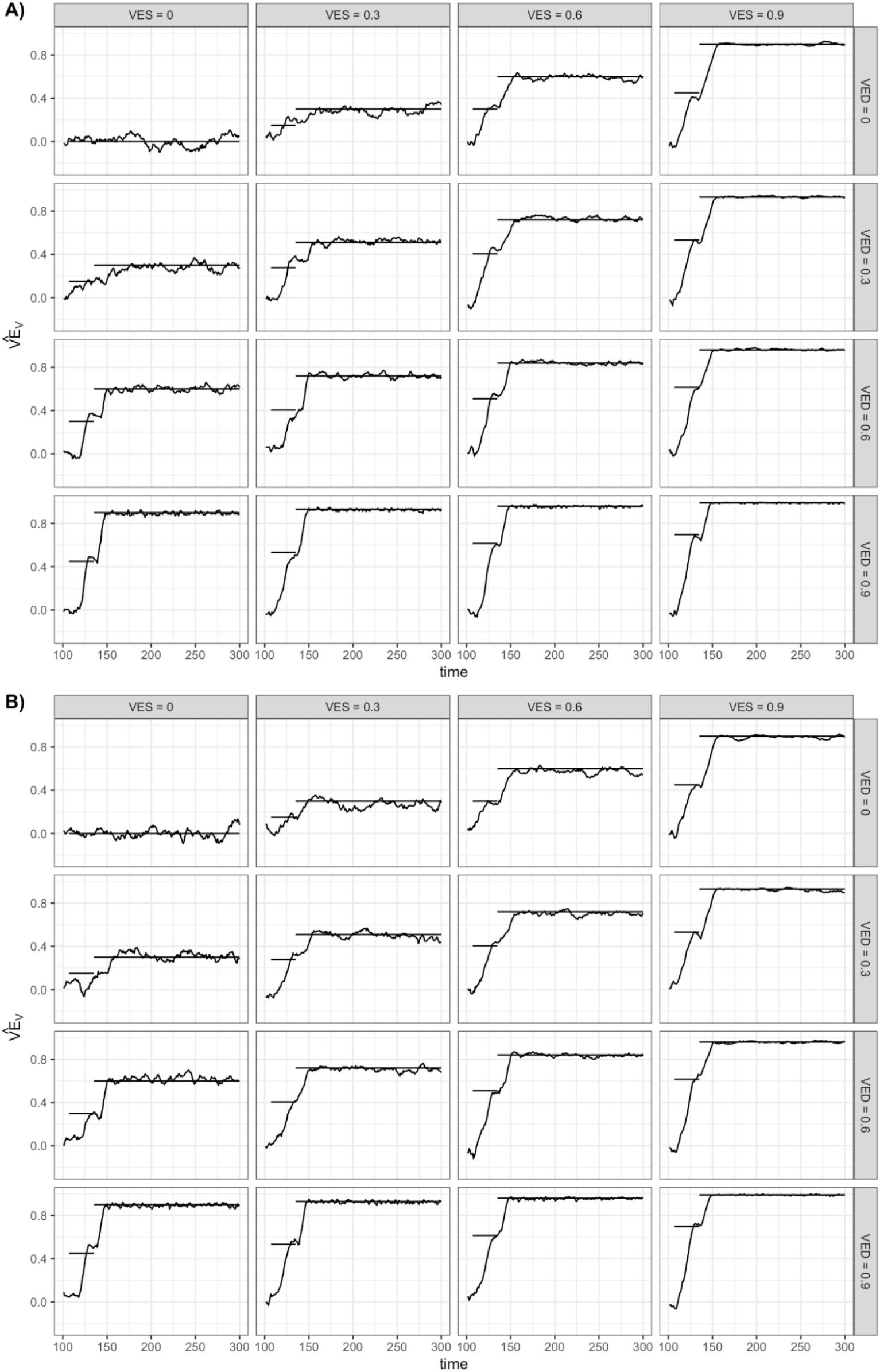
Vaccine efficacy for viral positivity. Results are shown of a 300-day simulation of a trial of 100,000 participants randomized 1:1 to vaccine or placebo on day 100 and exposed to a constant force of infection of 0.001 throughout the simulation. The different panels represent (left to right) simulations with *VE*_*S*_ = 0,0.3,0.6,0.9 and (top to bottom) *VE*_*D*_ = 0,0.3,0.6,0.9. We simulate a 2-dose regimen, 28 days apart with the first dose giving half the full efficacy and the effect of each dose starting one week after it is given, that is, on days 107 and 135 of the simulation. The solid black lines give the dose-1 and dose-2 predicted values for *VE*_*V*_ based on eq. 7, while the curves show the estimates obtained from the simulated data using eq. 6. Panel A shows the situation under the assumption that individuals naturally infected who recover (clear infection) become once again susceptible to reinfection (SEIS). Panel B makes the opposite assumption, that individuals naturally infected (whatever their vaccine status) are completely protected against reinfection for the duration of the simulation (SEIR).

In Fig. 1A, there is close agreement between the simulated curves and the predicted ones, after about day 150. During the first-dose period, estimated efficacy is noisy at the beginning but is typically below the predicted level, reflecting holdover of infections that occurred before (randomization+7 days), which are by assumption therefore unaffected by the vaccine. In contrast, toward the right-hand side of each panel, the agreement is nearly perfect apart from sampling error, because such holdover infections are vanishingly rare and the assumptions underlying eqs. 6 and 7 are met.

Fig. 1B shows a similar pattern, with the important exception that over time, as many individuals in the population are immune, the protection estimated from eq. 6 declines toward the null value. This is because both groups have fewer people at risk as immunity builds up, but when the vaccine has an effect, the placebo group is depleted of susceptible individuals faster than the vaccine group, rendering the two groups more similar and the apparent efficacy lower. This effect, which is a known complexity of randomized [27–30] and observational [31,32] studies of vaccine efficacy/effectiveness, is subtle in our primary analysis (Fig. 1B), but becomes more pronounced when there are longer times of follow up, higher forces of infection (Fig. S1), and greater heterogeneities in infection risk among the study population.

### Separate analyses of infections detected by testing those with symptoms and infections detected by testing cross sections of participants irrespective of symptoms improve interpretability of VE estimates

All trials of which we are aware for SARS-CoV-2 vaccines have had a primary endpoint of symptomatic disease, ascertained by asking every participant who experiences a defined profile of symptoms to get tested, and counting the outcome of COVID-19 when such a test is positive. As noted, some trials also test a subset of participants irrespective of symptoms, either at the visit for the second vaccine dose [2] or at defined intervals during follow up [16]. The primary endpoint measures vaccine efficacy against symptomatic infection, which has been called *VE*_*SP*_for vaccine efficacy against susceptibility or progression (that is, protection from symptomatic infection that could be preventing infection or preventing symptoms if an individual becomes infected), and is related to *VE*_*S*_ and *VE*_*P*_ (Table 1) by eq. 2 above.

Fig. 2 shows simulations similar to those above, but now with a virus assumed to cause symptoms in 1% (red) or 80% (blue) of infected individuals to demonstrate how the relationship between 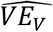 and 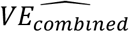 depends on the proportion symptomatic. In these simulations, all symptomatic individuals are assumed to be tested for the primary outcome on the day of symptom onset, and all asymptomatic individuals are not tested for the primary outcome. In addition, all individuals who have not yet experienced symptoms are tested for viral positivity and the combined VE is estimated. When only 1% of infected individuals are symptomatic, 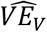 (solid) and 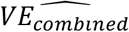 (dashed lines) are nearly identical. However, when 80% are symptomatic [33], 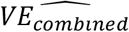 increases over time but falls below the expected *VE*_*SP*_, as it is a combination of VE against asymptomatic and symptomatic disease. If analysis is restricted to only non-symptomatic individuals (Fig. S2), when there is high *VE*_*P*_ (i.e. low *VE*_*S*_ for the same *VE*_*SP*_), 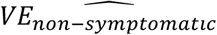 is lower than 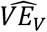.

**Figure 2.**
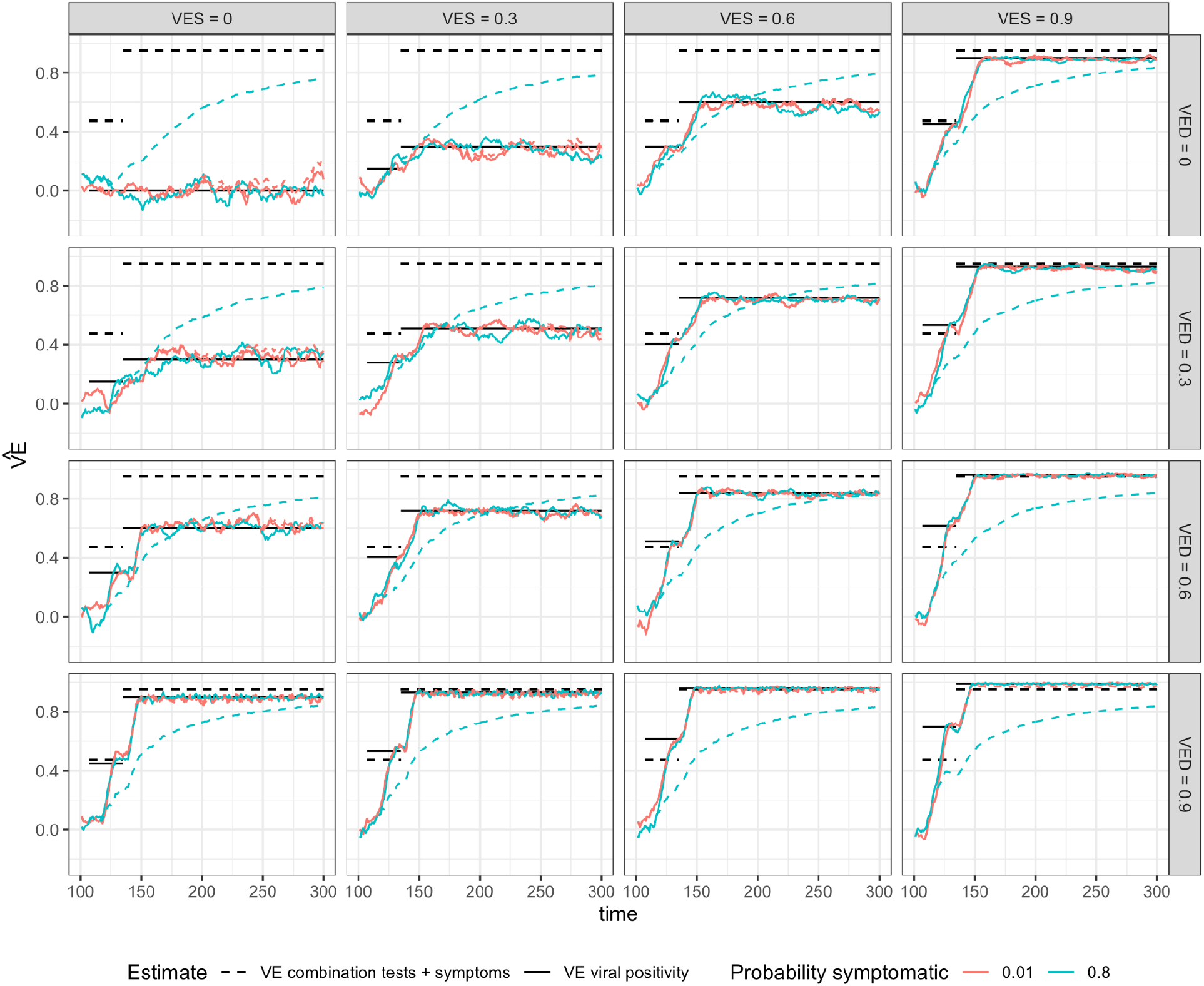
Vaccine efficacy for viral positivity and a combination of symptoms and testing. This figure shows the same simulations as Figure 1B with different analyses of the simulated data, comparing scenarios in which 1% and 80% of unvaccinated infections are symptomatic. The solid black lines give the dose-1 and dose-2 predicted values for *VE*_*V*_ based on eq. 7, while the solid curves show the estimates obtained from the simulated data using eq. 6 (the solid red line is the same as Figure 1B). The dashed lines give the dose-1 and dose-2 predicted values for *VE*_*SP*_, based on equation 2, while the dashed curves show the estimates of *VE*_*combined*_ obtained from the simulated data. When only 1% of infected individuals are symptomatic, the solid red and dashed red lines are nearly identical. However, when 80% are symptomatic [33], the dashed blue line increases over time but falls below the expected *VE*_*SP*_.

### Application to Moderna data

Table 2 shows data from the published RCT of the Moderna vaccine [2], in which participants returning for their second vaccine dose were tested by RT-PCR for SARS-CoV-2, with 39 and 15 testing positive without symptoms, respectively, in the placebo and vaccine group. If we assume that everyone in the modified intent-to-treat population not infected prior to the second dose was tested (this is not documented in the paper), this corresponds (Table 2) to an estimate of 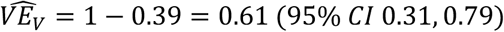, by eq. 6. Taken at face value, this implies that one dose reduces virus positivity by 61%; our simulations suggest this may be an underestimate for several reasons. First, this estimate only includes asymptomatic individuals. As Figure S2 shows, if only individuals not symptomatic at the time of swabbing were included, as in the Moderna study, there could be additional underestimation of 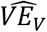 because vaccinated individuals without symptoms may disproportionately contribute to the non-symptomatic group. To resolve this potential bias, the data could be reanalyzed to also include anyone who tested positive on the day of the second dose and either was symptomatic or later became symptomatic. A modest underestimate could also occur due to holdover of individuals infected before the first dose took effect and still positive at the time of the second dose.

**Table 2.**

|  | Placebo | Vaccine |  |
| --- | --- | --- | --- |
| Positive | 39 | 15 |  |
| Approximate Inferred negative* | 14598-39-46=14513 | 14550-15-7=14528 |  |
| | | | $OR = (15)(14513)/[(39)(14528)] = 0.39$<br>95% CI: (0.21,0.69) |

\* Modified intent to treat population, minus those positive at the second vaccine visit (Table S18 of [2], minus those who became infected prior to the second dose (Fig 3 of [2])).

Finally, as noted above, one expects that *VE*_*V*_ ≤ *VE*_*T*_ (eq.8), so we conclude that the Moderna data from the second-dose swab provides evidence of at least a 61% (95% CI 31-79%) reduction in transmissibility due to a single dose of Moderna vaccine.

The VE estimate combining cases ascertained by symptoms and those ascertained by this testing protocol in Table S18 of [2] is 89.5% (85.1%-92.8%). As described above, this combined measure is an underestimate of the 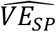 estimated in the study of 94.1% (89.3-96.8%).

## DISCUSSION

We have shown that if analyzed correctly, data from randomized trials that test a cross-section of vaccine and control recipients irrespective of symptoms on a given day for virus can estimate the vaccine efficacy against viral positivity. While a complete estimate of *VE*_*T*_ would require estimates of both *VE*_*V*_ and of the daily infectiousness of a vaccinated, infected individual compared to an unvaccinated, infected one, and their correlation across individuals, it is very likely in practice that *VE*_*V*_ is a lower bound on *VE*_*T*_: that is, an estimate from trial data of *VE*_*V*_ provides strong evidence that *VE*_*T*_ is at least as high.

Our main findings are as follows: first, that a single cross-sectional comparison of PCR positivity odds between individuals in vaccine vs. control groups provides a relatively accurate estimate, subject to sampling error, of vaccine effectiveness against viral positivity, which is a composite of effects in reducing susceptibility to infection and in reducing duration as described in Eq. 7. This can be shown analytically under certain assumptions. Second, we show by simulation that plausible deviations from these assumptions do not dramatically change results and, when they do, tend to bias toward the null hypothesis of no efficacy. A combined analysis of viral positivity detected due to symptoms and those detected by routine screening of non-symptomatic persons will be some combination of efficacy against viral positivity *VE*_*V*_ and against symptomatic infection *VE*_*SP*_ with no clear interpretation in terms of elementary quantities of interest. Thus separate analysis is recommended. Finally, if the cross-sectional sampling is restricted to those who are not symptomatic, it may underestimate *VE*_*V*_, especially for vaccines which are highly protective against symptoms (high *VE*_*P*_). We therefore recommend that the cross-sectional sample include those who are symptomatic. If this is infeasible (for example, if individuals are instructed not to come for a vaccine dose if they are symptomatic, and the testing happens at the vaccine dose), then we recommend that those who are tested because they are symptomatic and test positive on a particular day be included among the positives in the cross-section, constituting a partial exception to our recommendation of separate analyses.

Our results have been described in the setting of a randomized trial. These results apply also to observational studies as well insofar as they are designed to mimic a target trial [34] and achieve adequate control of confounding and other sources of bias.

In observational studies of vaccine effectiveness to date, cases have often been identified in whoever gets tested, for whatever reason [7,35,36]. These probably constitute a mix of (i) those tested because symptomatic, (ii) those tested because they are contacts of a known or suspected case (for example in a contact tracing investigation), and (iii) those tested without either reason, for example those who get tested in a regular program by their employer or those who get tested to comply with a travel restriction that requires a negative test before travel.

Those positive in group (i) are approximately equivalent, in the observational setting, to those who meet the primary outcome of confirmed COVID-19 from randomized trials. Those positive in group (iii) are perhaps equivalent, in the observational setting, to those who test positive in the routine follow-up of persons in a randomized trial. Group (ii) does not have a clear equivalent in the randomized trials, which typically do not gather information on contacts.

For observational studies, our results therefore imply that it would be ideal to analyze symptomatic cases separately from those routinely tested, and if possible to distinguish those tested due to possible exposure (group ii) from those tested for other reasons, such as for travel clearance (group iii). Those tested because they are symptomatic (group i) should be analyzed analogously to the trials, as the reduction in incidence rate. Those tested for exposure (group ii) are a group in which the efficacy measure is conditioned on exposure, and thus should be analyzed using methods to estimate the secondary attack rate, a risk measure. These recommendations follow standard approaches described in the landmark paper of Halloran et al. 1997 [37]. And those tested for neither reason (group iii) should be analyzed using the odds ratio approach described in this paper, extending others’ prior work [21].

We have not considered another approach that has been used in COVID-19 trials [19] to estimate the effect on asymptomatic infections: serologic testing of participants at the middle or end of the trial [30]. This can contribute to an estimate of *VE*_*S*_ and thus provide a lower bound on *VE*_*T*_, but does not address the duration of infectiousness or the viral shedding of the detected asymptomatic infection. Nevertheless, this is an important additional way to obtain evidence relevant to bounding the vaccine’s efficacy against transmission.

In summary, with careful analysis, data from swabs of individuals in vaccine and comparator arms can yield estimates of a key quantity, the vaccine’s efficacy in reducing viral positivity, likely a lower bound on the vaccine’s efficacy in reducing transmission. Future work should consider how quantitation of virus in both symptomatic and non-symptomatic individuals who do test positive may further refine these estimates.

## Data Availability

Code is available on github.

https://github.com/rek160/InterpretingVaccineEfficacy

## Acknowledgments

We thank Dr. Lee Kennedy-Shaffer for helpful discussion.

## Supplement

**Figure S1.**
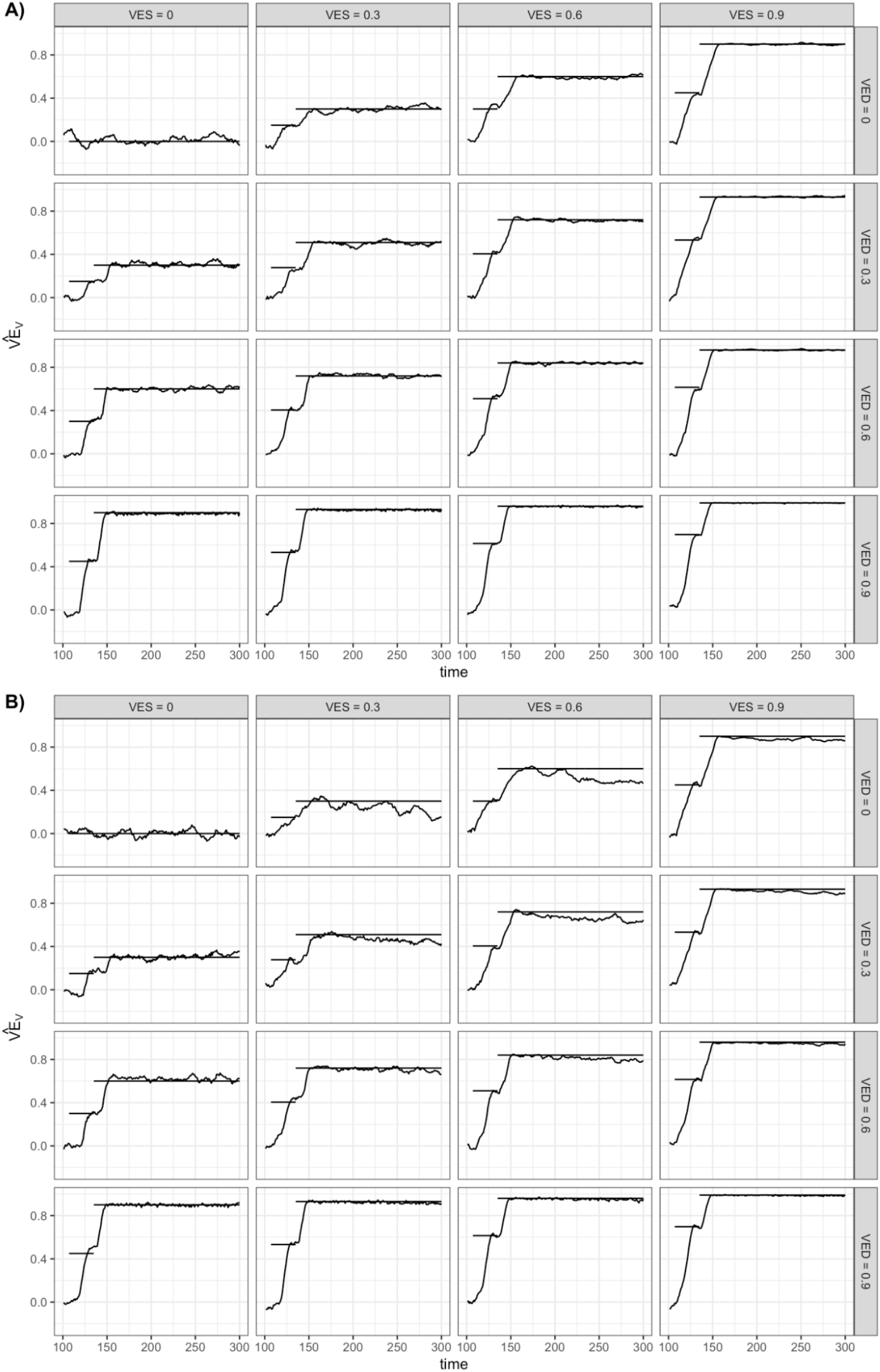
Vaccine efficacy for viral positivity with higher force of infection. This figure shows the same analysis as Figure 1 for a scenario with a constant force of infection of 0.003 (three times higher than the baseline scenario) throughout the simulation. The solid black lines give the dose-1 and dose-2 predicted values for *VE*_*v*_ based on eq. 7, while the curves show the estimates obtained from the simulated data using eq. 6. Panel A shows the situation under the assumption that individuals naturally infected who recover (clear infection) become once again susceptible to reinfection (SEIS). Panel B makes the opposite assumption, that individuals naturally infected (whatever their vaccine status) are completely protected against reinfection for the duration of the simulation (SEIR).

**Figure S2.**
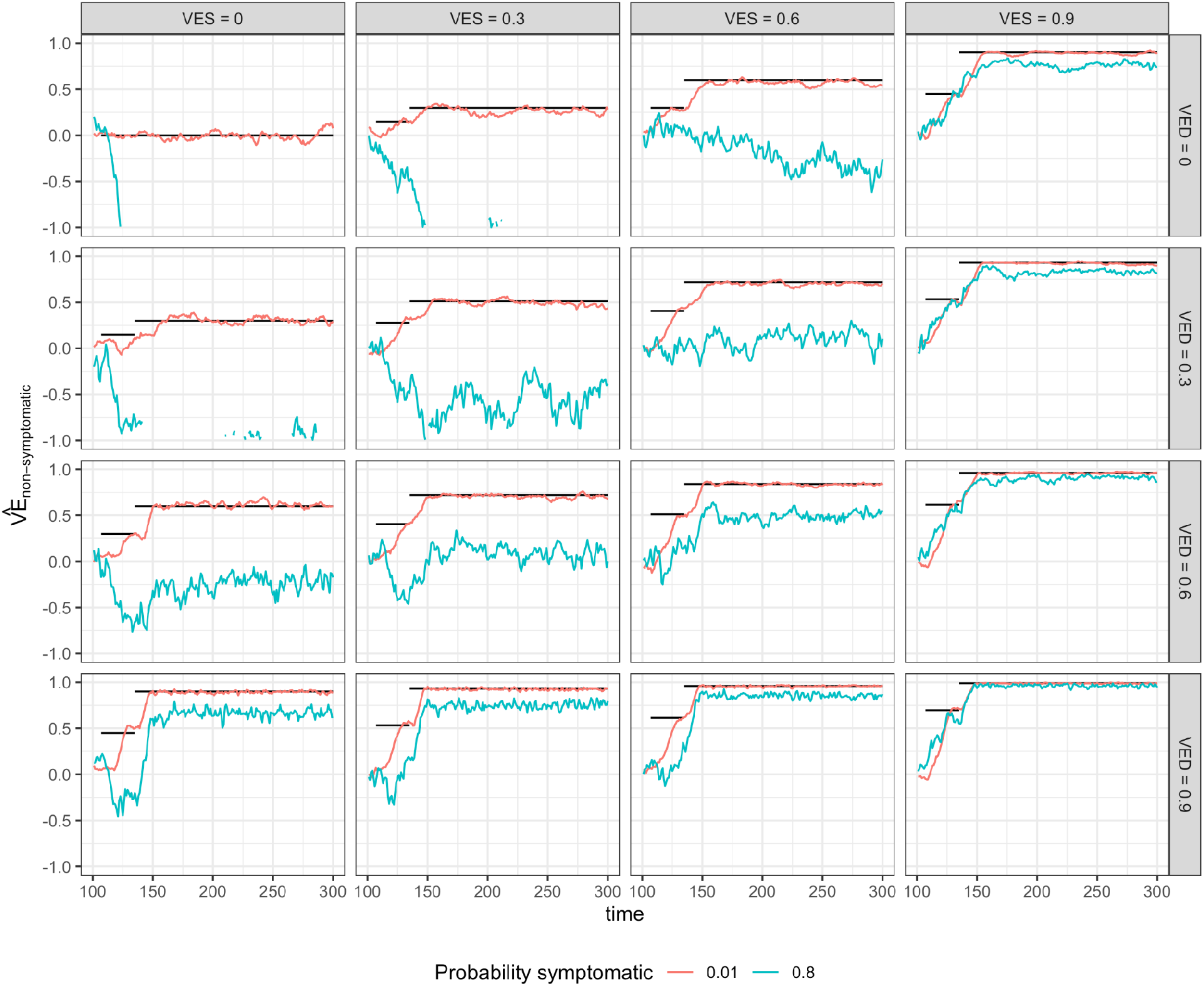
Vaccine efficacy for viral positivity in non-symptomatic participants. This figure shows the same simulations as Figure 2, with different analyses of the simulated data. The solid black lines give the dose-1 and dose-2 predicted values for *VE*_*V*_ based on eq. 7, while the curves show the estimates obtained from simulated estimates of *VE*_*non-symptomatic*_ Curves that “fall off” the bottom of the graph indicate efficacy estimates below -100%.

**Table S1.**

| Parameter | Value(s) |
| --- | --- |
| Day of enrollment in trial | 100 |
| Trial length (days) | 200 |
| Population size | 100,000 |
| Probability symptomatic | 0.80 [33], 0.01 |
| Latent period (days) | 3 [23] |
| Incubation period (days) | 5 [22] |
| Viral shedding period (days) | 15-21 [24,25] |
| Vaccine efficacy against susceptibility to infection | 0, 0.3, 0.6, 0.9 (half after first dose) |
| Vaccine efficacy against duration of infection | 0, 0.3, 0.6, 0.9 (half after first dose) |
| Vaccine efficacy against symptomatic infection | 0.95 (half after first dose) [1,2] |
| Time between 2 doses (days) | 28 [2] |
| Time for immune response to take effect (days) | 7 |
| Daily force of infection | 0.001, 0.003 |

